# The development of the Supplemental Nutrition Assistance Program enrollment accessibility (SNAP-Access) score

**DOI:** 10.1101/2022.03.09.22271551

**Authors:** Laura J. Samuel, Emily Xiao, Caroline Cerilli, Fiona Sweeney, Jessica Campanile, Nubaira Milki, Jared Smith, Jiafeng Zhu, Gayane Yenokyan, Adi Gherman, Varshini Varadaraj, Bonnielin K. Swenor

## Abstract

**Background:** The Supplemental Nutrition Assistance Program (SNAP) is a federally funded public benefit providing critical food assistance to millions of Americans. However, it is typically administered by states, creating potential variation in accessibility and transparency of information about enrollment for people with disabilities.

**Objective:** To create a method to assess the accessibility and transparency of information about the disability-inclusive process and practices of SNAP enrollment

**Methods:** Data was collected from SNAP landing and enrollment webpages from all 50 U.S. states, the District of Columbia, and New York City from June-August 2021. Based on key principles of universal design and accessibility, scores were determined for each SNAP program across three areas: flexibility in the enrollment process (6 points), efficiency of finding information about enrollment on SNAP websites (6 points), and the accessibility of SNAP webpages (6 points). Total scores were the sum of these sub-categories (18 points maximum).

**Results:** Of the 52 SNAP programs assessed, mean scores were 10.66 (SD=2.51) for the total score, 2.67 (SD=0.91) for flexibility in the enrollment process, 3.32 (SD=1.19) for efficiency of finding information about enrollment on SNAP websites, and 4.67 (SD=1.72) for the accessibility of SNAP webpage. A total of 0 SNAP programs received the maximum score (6 points) on flexibility, 2 programs the maximum on efficiency, and 31 programs the maximum on accessibility.

**Conclusions:** We found differences in the accessibility, flexibility, and efficiency of SNAP program enrollment information available on SNAP websites and outline much room for improvement across all three of these areas.

## Introduction

Adults with disabilities typically need more food assistance than their nondisabled peers but may experience more barriers to accessing food assistance programs, such as the Supplemental Nutrition Assistance Program (SNAP). Approximately 33% of households that include an adult with a disability are food insecure, compared with only 8% of households comprised of adults without disabilities.^1^ This is likely due to corresponding income inequality, as about 26% of adults with disabilities have incomes below the poverty limit, but only 11% of adults without disability fall below that income threshold.^2^

Enrollment in SNAP can reduce food insecurity ^3, 4^ and may ease the challenges faced by lower income adults to protect their health. As examples, SNAP participation is associated with less health care utilization,^5-7^ less cost-related medication non-adherence^8^ and, among those with diabetes, better glucose control.^9^ Despite these potential health gains, only 85% of eligible adults under 60 and less than half of eligible adults 60 years and older are enrolled in SNAP.^10^

Although many low-income individuals experience barriers to participating in SNAP, those with disabilities likely face additional challenges. Prior data indicate that the SNAP enrollment process can be cumbersome, and that there is stigma associated with using SNAP, which may discourage potential applicants.^11-13^ Although it seems likely that people with disabilities have even more barriers than their peers in navigating the complex enrollment and renewal processes, there has been little attention to defining those challenges or addressing them.

The established principles of universal design should be drawn upon to ensure equitable access to SNAP for all who are eligible, including those in the disability community.^14^ Key principles of universal design include accessibility, flexibility, and efficiency. Accessibility captures whether information is communicated effectively regardless of an individual’s disability status. Flexibility allows a wide range of formats to meet individual preferences and needs. Efficiency assesses whether processes within systems are simple, intuitive, and easy to navigate.^14^ Together, these principles can be used to evaluate whether systems and processes are inclusive of people with disabilities.

Food insecurity is experienced by one-third of households containing members with disabilities. Therefore, ensuring the SNAP program is accessible and inclusive is a public health priority. Although SNAP is a federally funded public benefit, it is typically administered by states, creating potential variation in SNAP program practices. Applicants typically rely on state agency websites as a first step in the SNAP enrollment process. The accessibility and transparency of information on these websites is a critical component of removing barriers to SNAP enrollment for people with disabilities. SNAP agencies should implement best practices for transparency of information on SNAP websites so that prospective applicants can easily find information about the program, determine eligibility, understand the application process, and find information about accommodations. However, currently there is no system to assess or track accessibility and disability inclusive practices of SNAP enrollment websites or processes.

This project’s aim was to develop a novel method to evaluate and compare the accessibility, flexibility, and efficiency of SNAP enrollment information available on SNAP websites. This information was disseminated via the Johns Hopkins Disability Health Research Center Supplemental Nutrition Assistance Program (SNAP) Disability Dashboard.^15^

## Methods and Analysis

From June through August of 2021, five trained student researchers conducted qualitative coding regarding the landing and enrollment pages of SNAP websites in all 50 U.S. states, the Washington District of Columbia (D.C.), and New York City. This study included people with disabilities and the sample included all SNAP programs in the U.S.

The SNAP landing page was defined as the introductory SNAP webpage for a given state, as linked on the USDA SNAP State Directory of Resources webpage. These landing pages generally included an overview of SNAP eligibility criteria, as well as links to SNAP enrollment applications, benefit utilization information, and related federal or state program webpages. In four cases where the website linked by the United States Department of Agriculture (USDA) directory was clearly incorrect (for example, if it led to an error message), the landing page was identified by inputting “[state name] SNAP website” into a search engine and finding the page that best matched the landing page. The enrollment pages requested an applicant’s name, address, and other identifying information, as well as information relevant to their household’s nutritional needs and socioeconomic status, for determination of SNAP benefits.

A data collection form was created in Microsoft Excel and pilot tested in an iterative process prior to data collection. Researchers asynchronously recorded information in unlinked documents prior to three consensus meetings. During consensus meetings, the researchers provided feedback and the team discussed improvements to the form. The final form collected data for 13 indicators related to flexibility and efficiency and identified the URL to be used for estimating accessibility based on SNAP program website data (**Supplemental Table 1)**. After data was extracted, quality reviews were conducted by one researcher for the 12 indicators of flexibility and efficiency among a randomly selected group of 10 states. Discrepancies were adjudicated by the first and senior author in a review meeting. This adjudication process resulted in changes for 14 values, or 12% of the 120 indicators reviewed across the 10 states (14 values/(10 states*12 indicators) = 12%).

### Flexibility

Flexibility indicates the number of modalities for SNAP enrollment to support access to a wide range of individuals. Scores were determined as the total number of the following modalities supported for enrollment in SNAP as described on the program website: in-person, online, email, mail-in, telephone and TTY. Programs were given 1.0 point or 0.5 points, where 1.0 point indicates greater flexibility of options, according to the rubric outlined in **Supplemental Table 1**.

### Efficiency

Efficiency scores determine how readily available SNAP enrollment information was on program websites. Scores were based on the following criteria: (1) if the SNAP program landing page had eligibility information or a direct link to an eligibility determination page, (2) if the program landing page had information on how to get accommodations for SNAP enrollment, (3) if a telephone help line was provided on the program website, (4) if there was a TTY help line listed on the program website, (5) if the program landing page had links to an online enrollment form and/or a PDF enrollment form, and (6) if the program landing page had links to a large print PDF version of the SNAP enrollment form.

### Accessibility

Accessibility scores were calculated using an average of automated accessibility test data across a broad sample of SNAP site pages and high-level manual accessibility testing on both the SNAP information and registration pages, when available. The WAVE accessibility tool ^16^ was used to collect automatic accessibility data. Each site was assigned an automated accessibility score based on the number of detected errors, the density of errors (number of errors by number of web page elements), and number of likely or potential accessibility errors. These data were normalized to the WebAIM Million sample of one million homepages ^17^ to indicate how the tested web site compares to web pages generally across the web. The automated scores for SNAP pages were typically higher than other collected scores, but this is primarily because the average web page has significant accessibility barriers and SNAP pages fared only slightly better than average. Expert testers then evaluated 10 aspects of web accessibility on the landing and enrollment pages, assigning values based on potential impact of accessibility issues on individuals with disabilities. The 10 aspects of web accessibility included (1) accuracy of the document’s defined language, (2) appropriateness of image alternative text, (3) impact of empty links and buttons, (4) impact of labeled or unlabeled form inputs, (5) low contrast content, (6) appropriateness of page title, (7) animation and movement, (8) keyboard focus indicators, (9) keyboard accessibility, and (10) page reflow/responsiveness. The overall accessibility values were the average of the automated scores and manual scores for each program. The final accessibility scores were then based on tertiles of these total values, as follows: the lowest tertile (overall accessibility values below 4.3) received 0 points, the middle tertile (overall accessibility values at or above 4.3 and below 7.0) received 3 points, and the highest tertile (overall accessibility values of 7.0 or above) received 6 points.

Total SNAP scores were calculated as the sum of the flexibility, efficiency, and accessibility scores for each program. A maximum of 18 total points were possible.

### Statistical Methods

Summary statistics such as mean, standard deviation, and range were provided for the total score, as well as each subcategory (flexibility, efficiency, and accessibility). Total and sub-category scores were each divided into tertiles: lowest, middle, and highest scores. Using this data, heat maps of the U.S. were created to represent the geographical distribution of these score tertiles (**Figures 1 to 4**). Statistical analyses were completed using R statistical software (version 4.1.1).

**Figure 1.**
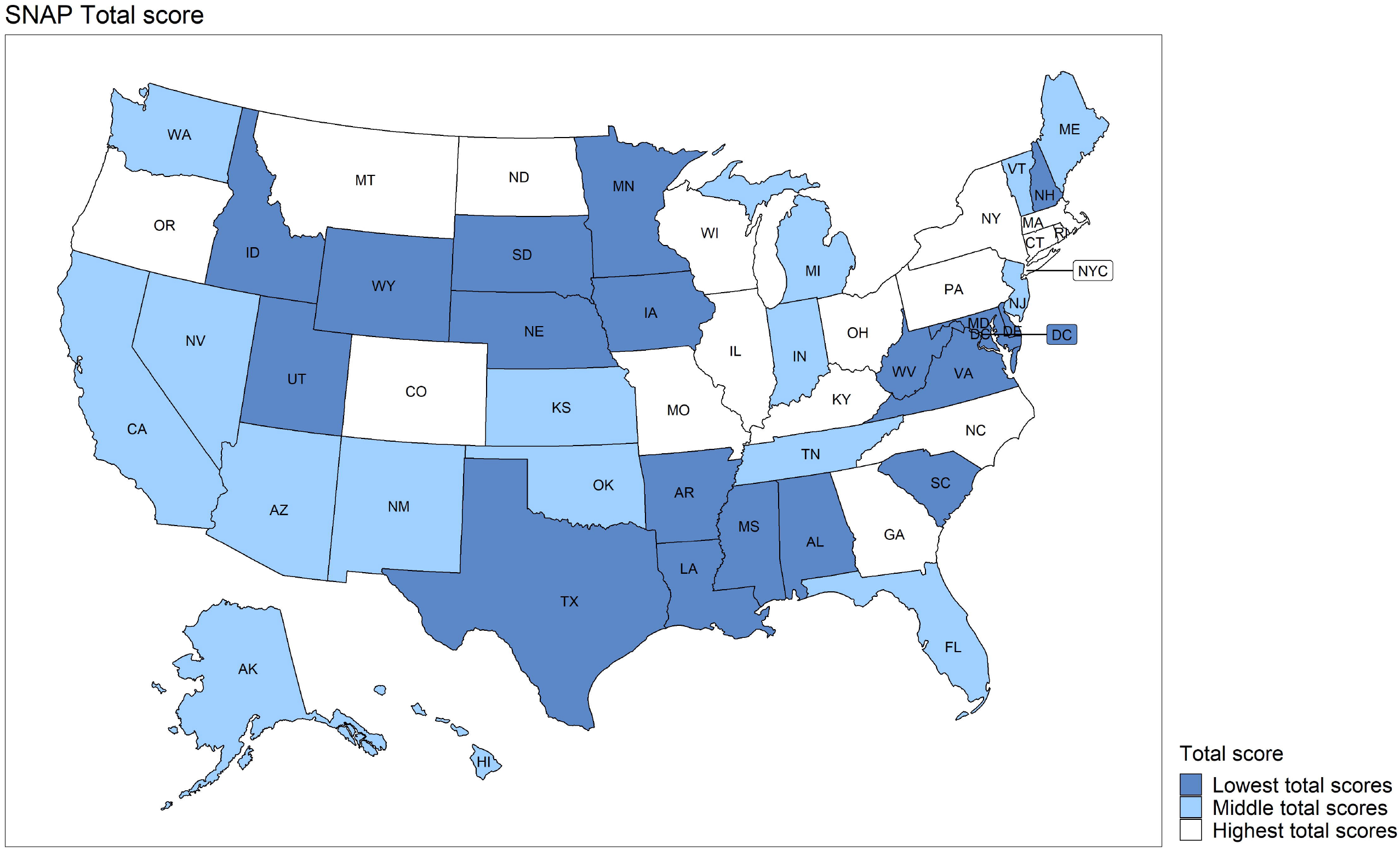
Supplemental Nutrition Assistance Program (SNAP) Enrollment Accessibility and Disability Inclusion: Total Scores across the United States, 2021

**Figure 2.**
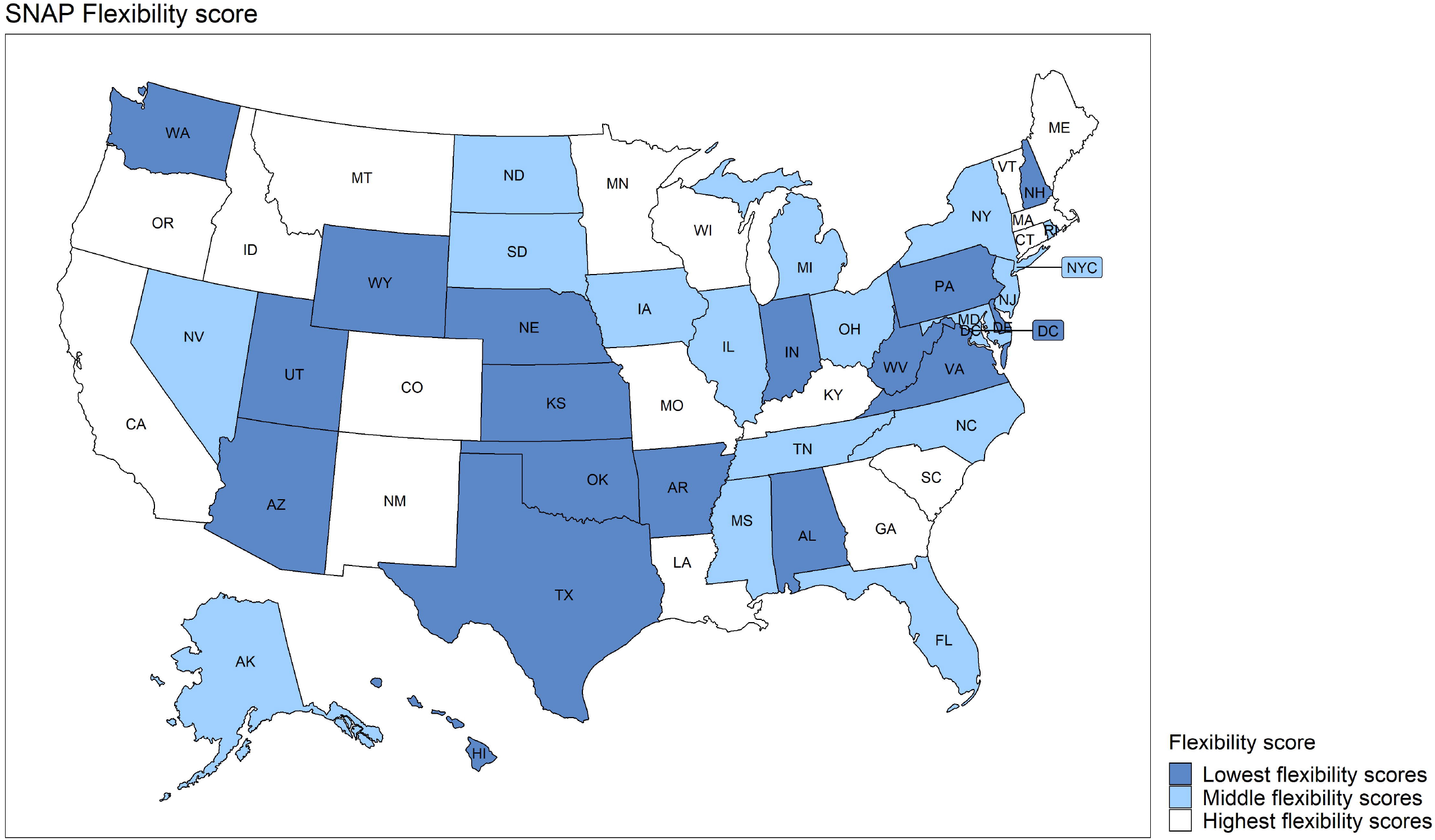
Flexibility in Supplemental Nutrition Assistance Program (SNAP) Enrollment Total Scores across the United States, 2021

**Figure 3.**
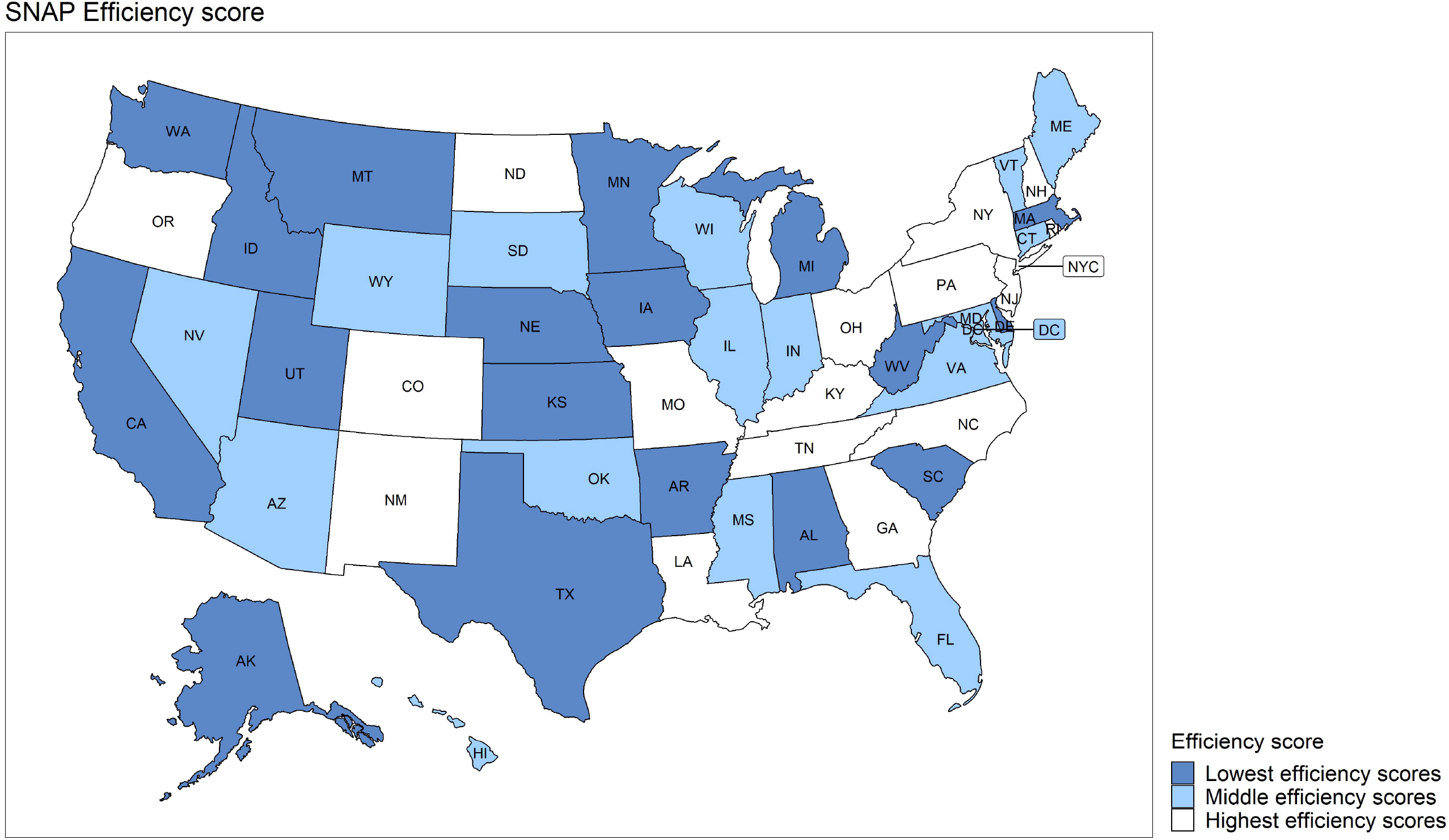
Efficiency of Supplemental Nutrition Assistance Program (SNAP) Enrollment Total Scores across the United States, 2021

**Figure 4.**
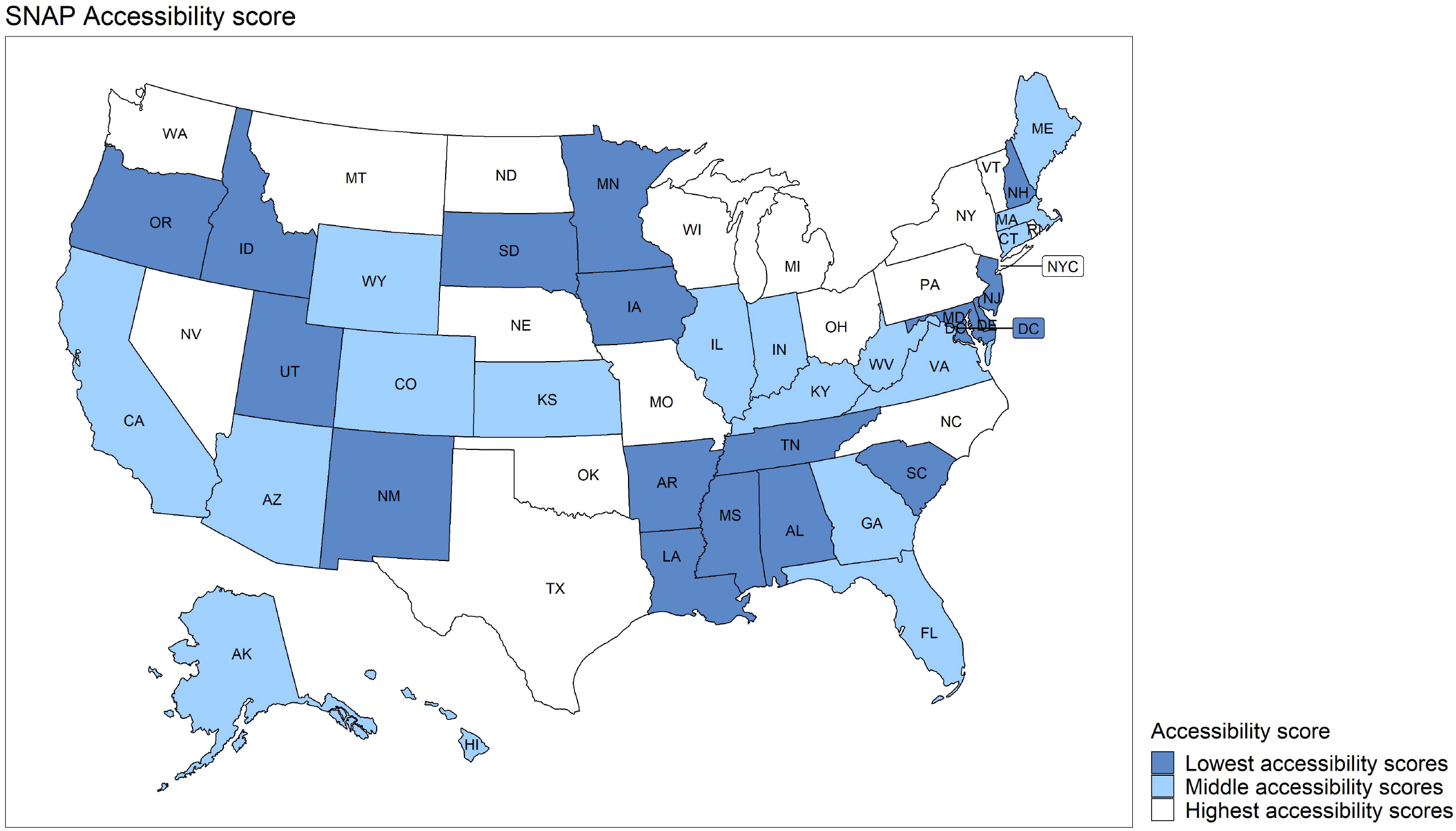
Accessibility of Supplemental Nutrition Assistance Program (SNAP) Websites Total Scores across the United States, 2021

## Results

Of the 52 SNAP programs assessed, which included programs in all 50 states, the Washington D.C. and New York City, the mean total score was 10.66 (SD=2.51) out of 18 possible total points and ranged from 5.5 to 16.5. The mean scores were 2.67 (SD=0.91) for *flexibility of SNAP enrollment* scores, 3.32 (SD=1.19) for *efficiency of SNAP enrollment*, and 4.67 (SD=1.72) for a*ccessibility* (**Table 1**). In the flexibility of SNAP enrollment category, the majority of programs had enrollment options described on the websites for in-person (85%), online (92%), and mail-in (75%) options, but not for the email (8%), telephone (25%), and TTY (10%) options. In the efficiency of SNAP enrollment category, the majority of programs listed eligibility information (85%), a telephone help line (85%), and an online enrollment form linked to state (94%), but not enrollment accommodation information (31%), TTY help line (46%), or a large-print PDF enrollment form (12%). Scores ranged across SNAP programs from 1 to 5 for the flexibility sub-category, 1.5 to 6 for efficiency, and 0 to 6 for accessibility (**Table 2**). The number of SNAP programs that received the maximum score (6 points) within each sub-category differed: 0 SNAP programs for flexibility, 2 programs for efficiency, and 31 programs for accessibility. There were differences in these scores when examined by tertile ranking across the U.S. (**Figures 1 to 4**).

**Table 1.** Supplemental Nutrition Assistance Program (SNAP) for program characteristics in all 50 U.S. states, Washington D.C. and New York City, 2021

|  |  |  |  |
| --- | --- | --- | --- |
|  | SNAP program characteristics |  |  |
| Flexibility of SNAP Enrollment |  |  |  |
| Mean enrollment score of SNAP websites (SD) | 2.67 (0.91) |  |  |
| Enrollment options described on website (%) | No, n (%) | Yes, only by jurisdiction, n (%) | Yes, for the state, n (%) |
| In-person<sup>a</sup> | 8 (15%) | 0 (0%) | 44 (85%) |
| Online | 4 (8%) | 0 (0%) | 48 (92%) |
| Email | 48 (92%) | 0 (0%) | 4 (8%) |
| Mail in | 13 (25%) | 26 (50%) | 13 (25%) |
| Telephone | 39 (75%) | 2 (4%) | 11 (21%) |
| TTY | 47 (90%) | 0 (0%) | 5 (10%) |
| Efficiency of SNAP Enrollment |  |  |  |
| Mean Efficiency score of SNAP websites (SD) | 3.32 (1.19) |  |  |
| Efficiency of SNAP websites (%) | No, n (%) | Yes, but not clearly indicated, n (%) | Yes, and clearly indicated, n (%) |
| Has eligibility information | 8 (15%) | N/A | 44 (85%) |
| Has enrollment accommodation information | 36 (69%) | 0 (0%) | 16 (31%) |
| Has a telephone help line | 8 (15%) | 6 (12%) | 38 (73%) |
| Has a TTY help line | 28 (54%) | 0 (0%) | 24 (46%) |
| Has online enrollment form linked to state | 3 (6%) | 15 (29%) | 34 (65%) |
| Has large-print PDF enrollment form | 46 (88%) | 0 (0%) | 6 (12%) |
| **Accessibility of SNAP Enrollment** |  |  |  |
| Mean accessibility score of SNAP websites (SD) | 4.67 (1.72) |  |  |

\* For most enrollment options, state-wide services were the preferred option, except that by jurisdiction was preferred for in-person enrollment.

**Table 2.** Supplemental Nutrition Assistance Program (SNAP) enrollment accessibility and disability inclusion total scores in all 50 U.S. states, Washington D.C. and New York City

| State | Total Score<br>(18 pts max) | Flexibility in SNAP<br>Enrollment Score<br>(6 pts max) | Efficiency of SNAP<br>Enrollment Score<br>(6 pts max) | SNAP Website<br>Accessibility Score<br>(6 pts max) |
| --- | --- | --- | --- | --- |
| Alabama | 8.0 | 2.5 | 2.5 | 3.0 |
| Alaska | 11.0 | 3.0 | 2.0 | 6.0 |
| Arizona | 11.5 | 2.5 | 3.0 | 6.0 |
| Arkansas | 7.5 | 2.5 | 2.0 | 3.0 |
| California | 12.5 | 4.0 | 2.5 | 6.0 |
| Colorado | 14.5 | 3.5 | 5.0 | 6.0 |
| Connecticut | 14.0 | 4.0 | 4.0 | 6.0 |
| Delaware | 8.0 | 2.5 | 2.5 | 3.0 |
| District of Columbia | 9.5 | 2.5 | 4.0 | 3.0 |
| Florida | 12.5 | 3.0 | 3.5 | 6.0 |
| Georgia | 16.5 | 4.5 | 6.0 | 6.0 |
| Hawaii | 11.5 | 2.5 | 3.0 | 6.0 |
| Idaho | 8.5 | 4.0 | 1.5 | 3.0 |
| Illinois | 12.5 | 2.5 | 4.0 | 6.0 |
| Indiana | 10.5 | 1.0 | 3.5 | 6.0 |
| Iowa | 7.5 | 2.5 | 2.0 | 3.0 |
| Kansas | 9.5 | 1.0 | 2.5 | 6.0 |
| Kentucky | 15.0 | 4.0 | 5.0 | 6.0 |
| Louisiana | 9.0 | 5.0 | 4.0 | 0.0 |
| Maine | 12.0 | 3.0 | 3.0 | 6.0 |
| Maryland | 9.0 | 2.5 | 3.5 | 3.0 |
| Massachusetts | 12.5 | 4.0 | 2.5 | 6.0 |
| Michigan | 11.0 | 2.5 | 2.5 | 6.0 |
| Minnesota | 9.0 | 3.5 | 2.5 | 3.0 |
| Mississippi | 8.5 | 2.5 | 3.0 | 3.0 |
| Missouri | 14.0 | 3.0 | 5.0 | 6.0 |
| Montana | 12.5 | 4.0 | 2.5 | 6.0 |
| Nebraska | 9.0 | 1.0 | 2.0 | 6.0 |
| Nevada | 11.0 | 2.5 | 2.5 | 6.0 |
| New Hampshire | 8.5 | 1.5 | 4.0 | 3.0 |
| New Jersey | 9.5 | 2.5 | 4.0 | 3.0 |
| New Mexico | 10.0 | 3.0 | 4.0 | 3.0 |
| New York | 13.5 | 2.5 | 5.0 | 6.0 |
| New York City | 13.5 | 2.5 | 5.0 | 6.0 |
| North Carolina | 13.5 | 2.5 | 5.0 | 6.0 |
| North Dakota | 12.5 | 2.5 | 4.0 | 6.0 |
| Ohio | 12.5 | 2.5 | 4.0 | 6.0 |
| Oklahoma | 11.0 | 2.0 | 3.0 | 6.0 |
| Oregon | 12.5 | 3.5 | 6.0 | 3.0 |
| Pennsylvania | 13.0 | 2.0 | 5.0 | 6.0 |
| Rhode Island | 12.5 | 2.5 | 4.0 | 6.0 |
| South Carolina | 8.0 | 3.0 | 2.0 | 3.0 |
| South Dakota | 5.5 | 2.5 | 3.0 | 0.0 |
| Tennessee | 10.5 | 2.5 | 5.0 | 3.0 |
| Texas | 8.5 | 1.0 | 1.5 | 6.0 |
| Utah | 7.0 | 2.0 | 2.0 | 3.0 |
| Vermont | 11.5 | 3.0 | 2.5 | 6.0 |
| Virginia | 8.0 | 2.0 | 3.0 | 3.0 |
| Washington | 10.0 | 2.0 | 2.0 | 6.0 |
| West Virginia | 5.5 | 1.0 | 1.5 | 3.0 |
| Wisconsin | 12.5 | 3.5 | 3.0 | 6.0 |
| Wyoming | 7.0 | 1.5 | 2.5 | 3.0 |

## Discussion

This project found differences in the accessibility, flexibility and efficiency of SNAP program enrollment information available on SNAP websites and outline much room for improvement across all three of these areas. Together with evidence showing disproportionate food insecurity burden among people with disabilities,^1^ these results suggest that people with disabilities are doubly disadvantaged with regard to food access. They are not only more in need of food assistance than their peers, but also face more barriers in the information gathering and enrollment process for food assistance programs.

These results are among the first to quantify the accessibility of the SNAP enrollment process. Although these results identify important barriers to the SNAP program for people with disabilities, this study focused on the tasks required to prepare a SNAP application, which is only the first step of the SNAP enrollment process. There are many steps to enrollment, utilization, and recertification that were not examined in this study, including submitting applications with complete documentation, completing an interview, utilizing benefits, updating addresses and other household information as needed, and recertification. All these critical steps should also be examined regarding inclusiveness for people with disabilities. In addition, similar work is needed to understand inclusion and accessibility across all agencies in the social service sector related to housing, unemployment benefits, health insurance, tax credits, and income supplements.

These results have important implications for policies, processes and practices for public benefits such as SNAP by encouraging 1) a system of accountability for inclusion, 2) state partnerships with disability communities, and 3) federal funding to support these efforts. First, there is a need for a better system of accountability to ensure inclusion of people with disabilities in federally-funded benefit programs such as SNAP. Section 504 of the Rehabilitation Act of 1973 protects people with disabilities from being “excluded from the participation in, be denied the benefits of, or be subjected to discrimination under any program or activity receiving federal financial assistance”, including programs such as SNAP. Under Title II of the Americans with Disabilities Act (ADA) which applies to state and local government entities, subtitle A, qualified individuals with disabilities are protected from discrimination on the basis of disability in services, programs, and activities provided by state and local government entities. This includes ensuring processes do not screen out or prohibit qualified individuals with disabilities from accessing or enrolling in services. Additionally, both the ADA and Rehabilitation Act include protections against inaccessibility of information, including online information. However, the current paradigm of these disability rights laws puts the onus of compliance on the individual, as it usually requires the person with a disability who may experience gaps in accessibility of the SNAP enrollment process to file a complaint to be afforded remediation of those gaps. This dashboard intends to build a data infrastructure to change that paradigm. By collecting and sharing data on SNAP enrollment information accessibility, flexibility, and efficiency, we aim to support the advancement of disability inclusion in SNAP.

Second, state leadership must partner with disability communities to develop inclusive social services. For state practices, processes, and policies to better serve and include the disability community, people with disabilities must be included as advisors, leaders, and staff at agencies that provide social services. Further, there must be clear lines of communication between these state agencies and disability communities, including transparent processes for sharing grievances or concerns with the current systems. Disability input and inclusion is not a one-time effort. As technology evolves, and programs change, input from disability communities is needed.

Third, federal funding is needed to assist states in becoming more accessible and inclusive of people with disabilities. Funding should be commensurate upon ensuring processes and practices stay up to date with best practices and new technology, and address issues that are raised by those in the disability community. Including additional programmatic supports and requirements for state SNAP budgets can help make this program accessible and equitable for people with disabilities.

Importantly, states should take action to make the SNAP enrollment process more efficient. This includes identifying and eliminating inefficient steps of the enrollment and recertification process for people with disabilities. For example, the Combined Application Project^18^ allows SNAP programs to partner with the Social Security Administration so that people who are already known to be low-income because they receive Supplemental Security Income (SSI) can enroll in SNAP either automatically or with dramatically reduced application requirements. Because 86% of SSI recipients also have a disability, this can both improve inclusion for beneficiaries and reduce administrative burden for SNAP programs. Yet, despite evidence that the Combined Application Project can increase SNAP participation,^18^ it still has not been implemented in 32 states and Washington, D.C. Similarly, the Elderly Simplified Application Project can reduce the documentation requirements for older adults with disabilities and can waive the recertification interview requirement for individuals who rely on income sources such as Social Security that are fixed over time. Finally, allowing broad-based categorical eligibility can simplify the SNAP enrollment process by raising the income eligibility limit and/or eliminating the asset test. The asset test is a practice intended to limit SNAP eligibility to households with the smallest amounts of saved money or vehicle assets but individuals with disabilities are likely more reliant on assets than their peers without disabilities because of limitations in earning wages or income. In addition, there is good evidence that individuals with disabilities need 28% more income to maintain the same standard of living as a household of the same size where no one has a disability due to extra costs for day-to-day needs like food preparation or transportation.^19^ Therefore, use of broad-based categorical eligibility to both eliminate the asset test and increase the income limit for people with disabilities may level the playing field to ensure their food needs are met.

In addition to eliminating burdensome steps in the enrollment process, SNAP programs could improve disability inclusion by providing more flexibility, assistance and accommodations with the enrollment process. This dashboard provides the infrastructure for SNAP programs to compare and learn from best practices in other states, and implement new enrollment modalities. In addition, states can leverage administrative data from Medicaid, Low Income Home Energy Assistance Program, and SNAP programs to identify income-eligible individuals who are not enrolled in SNAP and conduct targeted outreach to increase participation. Cross-benefit enrollment interventions have been shown to increase SNAP participation^20^ and may be particularly important for individuals with disabilities who may face disproportionate challenges navigating the complex enrollment processes.

## Limitations & Strengths

Results are limited to information obtained from websites; there may be additional enrollment modalities beyond those described on the website and accommodations may be offered after an individual initiates an online application or downloads a PDF form. However, based on the principle of efficiency, we believe it is important to provide that information before initiating applications so that individuals with disabilities have the opportunity to make an informed enrollment plan based on the modalities and accommodations that would work best for them. Strengths of this study include having a conceptual grounding in key principles of universal design and inclusion of SNAP programs across all 50 states, Washington D.C. and New York City.

## Conclusions

This dashboard shows gaps in ensuring that individuals with disabilities can access SNAP enrollment websites and online information about the SNAP enrollment process. There is a need to create best practices that improve accessibility and increase SNAP enrollment, additional federal support, and improved input from the disability community.

## Data Availability

All data produced are available online at https://disabilityhealth.jhu.edu/snapdashboard/

