## Supplemental Table 1 for "The development of the Supplemental Nutrition Assistance Program enrollment accessibility (SNAP-Access) score"

**Supplemental Table 1**. Supplemental Nutrition Assistance Program (SNAP) program website data scoring tool

| **Indicator** | **Score** | **Description** | **Examples** |
| --- | --- | --- | --- |
| **FLEXIBILITY IN ENROLLMENT, 6 total points** | | | |
| In-person enrollment | 1 pt | In-person enrollment is available by jurisdiction (i.e., applicant is directed to nearest county office). The SNAP landing or enrollment page provides (1) physical addresses or (2) a link to a local office directory. | Illinois has an enrollment landing page that lists options for applying. One of these is to "carry, mail, or fax it to your local Family Community Research Center" and links directly to its DHS Office Locator. |
|  | 0.5 pt | In-person enrollment is available state-wide, and the address is provided on the SNAP landing or enrollment webpage. |  |
|  | 0 pt | In-person enrollment is not listed as an option on the SNAP landing or enrollment page. | Maine and Missouri are examples of states that clearly list ways to apply, but in-person enrollment is not included. |
| Online enrollment | 1 pt | Online enrollment via a state-wide portal is listed as an option on the SNAP landing or enrollment page. | Connecticut's "Apply" page (linked in sidebar on landing page) states as one option: "To apply online, please visit [www.connect.ct.gov](http://www.connect.ct.gov/), under 'Apply for Benefits.'" |
|  | 0.5 pt | Online enrollment is listed as an option by jurisdiction, and the SNAP landing or enrollment page provides relevant links or a directory. |  |
|  | 0 pt | Online enrollment is not listed as an option on the SNAP landing or enrollment page. | Idaho does not offer an online enrollment option. |
| Email enrollment | 1 pt | Email enrollment is listed as an option, and a state-wide email address is provided on the SNAP landing or enrollment page. | Oregon lists options on its enrollment page for submitting an application. These include emailing the application to the address provided. |
|  | 0.5 pt | Email enrollment is listed as an option by jurisdiction, and the SNAP landing or enrollment page provides (1) email addresses or (2) a link to a local office directory. |  |
|  | 0 pt | Email enrollment is not listed as an option on the SNAP landing or enrollment page. | Alabama indicates in its PDF application that the form can be emailed to the local Food Assistance Office, but this information is not found on the actual SNAP webpage. |
| Mail enrollment | 1 pt | Mail-in enrollment is listed as an option, and a state-wide mailing address is provided on the SNAP landing or enrollment page. | Louisiana lists options on its enrollment page for submitting a paper application. One option is to mail the completed form to the state-wide Document Processing Center (address provided). |
|  | 0.5 pt | Mail-in enrollment is listed as an option by jurisdiction, and the SNAP landing or enrollment page provides (1) mailing addresses or (2) a link to a local office directory. | Michigan states that applications can be mailed to a local office. A link to an office directory is provided on the same page. |
|  | 0 pt | Mail-in enrollment is not listed as an option on the SNAP landing or enrollment page. | Pennsylvania does not include an option for mail-in enrollment. |
| Telephone enrollment | 1 pt | Telephone enrollment is listed as an option, and a state-wide phone number is found on the SNAP landing or enrollment page. | Washington indicates on its landing page that an application can be completed over the phone (phone number provided). |
|  | 0.5 pt | Telephone enrollment is listed as an option by jurisdiction, and the SNAP landing or enrollment page provides (1) phone numbers or (2) a link to a local office directory. | Wisconsin provides a directory for identifying the contact information for the applicant's local office. |
|  | 0 pt | Telephone enrollment is not listed as an option on the SNAP landing or enrollment page. | South Carolina and many other states do not list telephone enrollment as an option. |
| TTY enrollment | 1 pt | A state-wide TTY option is provided for enrollment on the SNAP landing or enrollment page. | Kentucky offers both a phone number for telephone enrollment and an accompanying TTY number. |
|  | 0.5 pt | TTY numbers for enrollment are provided by jurisdiction. The SNAP landing or enrollment page provides these TTY numbers, or a local directory that contains TTY numbers. |  |
|  | 0 pt | A TTY option is not listed for telephone enrollment, or telephone enrollment is not listed as an option on the SNAP landing or enrollment page. | Massachusetts offers telephone enrollment via its DTA Assistance Line, but no TTY number is provided. |
| **EFFICIENCY OF ENROLLMENT, 6 points total** | | | |
| Are there options to get accommodations for SNAP enrollment? | 1 pt | The SNAP landing page, or any directly linked page, includes specific instructions for what accommodations are available and how to request them. | Michigan's website: "DHS can assist you if you need help filling out the application or need someone to read the application to you. [...] If you are unable to come into the office to complete an application because of a disability, you may contact your local MDHHS Office to request that someone come to your home to help you complete one." |
|  | 0.5 pt | The SNAP webpage says that accommodations are available, but does not state on the landing page or enrollment page what these accommodations are or how to obtain them. |  |
|  | 0 pt | The SNAP landing or enrollment page has no language about available accommodations or avenues for obtaining them. | South Dakota's PDF application allows applicant to indicate whether they are "Visual or Hearing Impaired" and in need of free interpreter services. This information does not receive credit because it was not available on the landing page or linked webpages. |
| Is there a help line? | 1 pt | Phone number is provided on landing page or linked webpages. It is (1) clearly indicated as SNAP-specific help line, or (2) a state-wide/consolidated contact line that lists SNAP as one of its services. | Georgia provides a telephone number for its "SNAP Customer Contact Center." |
|  | 0.5 pt | Phone number is provided on landing page or linked webpages, but is not clearly indicated as a SNAP-specific help line. This still receives partial credit if it is indicated as a customer-facing call line. | Kansas' Economic and Employment Services program provides a "Customer Service" phone number under its "Program Contacts" page. |
|  | 0 pt | Phone number cannot be found on landing page or linked webpages. A state-wide departmental phone number receives no credit if it is neither indicated as a customer help line, nor indicated for questions about SNAP. | Texas does not provide any contact information. |
| Is there a help line TTY? | 1 pt | A TTY help option is provided on landing page or linked webpages. It is (1) clearly indicated as SNAP-specific help line, or (2) a state-wide/consolidated contact line that lists SNAP as one of its services. | New Hampshire provides a TTY number for its SNAP customer line. |
|  | 0.5 pt | A TTY help line is provided on landing page or linked webpages without an indication that it is SNAP-specific. It receives partial credit if it is indicated as a customer-facing help line. |  |
|  | 0 pt | A TTY help option is not provided on the landing page or linked webpages. | Virginia has a SNAP hotline but does not provide a TTY number. |
| Does state SNAP landing page have information about eligibility or a direct link to an eligibility page? | 1 pt | Eligibility information or criteria can be found on the landing page or within one click (i.e., a clearly indicated direct link). | Colorado includes a "What you need to know" section in its landing page, with one of the drop-down questions being "Who is eligible for SNAP?" |
|  | 0 pt | Finding information about eligibility requires more than one click away from the landing page. | Idaho's landing page does not include text about eligibility, nor does it have a clear link to an eligibility page. |
| Is enrollment form directly linked to state SNAP landing page? | 1 pt | Direct links to both the online and PDF applications are found on the landing page (only 1 click required). If the applicant must navigate to another page (2 clicks required), credit is still given if the intermediate link is conspicuously indicated as a path to apply (e.g., a large button or sidebar link with text "How to Apply"). | Hawaii provides direct and clearly worded links on its landing page. |
|  | 0.5 pt | Only one of the two options (online or PDF) can be reached with the above method. | Wisconsin has both an online and PDF application, but only the online form is accessible in the described way. |
|  | 0 pt | Neither form can be reached with the above method. | Rhode Island requires more than two clicks. Clicking "Eligibility & How to Apply" in the SNAP-specific sidebar directs applicants to a page where they must scroll down and find another link for options to "Apply for DHS Benefits," and the forms are found here. |
| Large print form availability? | 1 pt | A large print application form is available and can be accessed within 2 clicks from the landing page (see method above). | Georgia includes a large print form in its bulleted list of application options on its landing page. |
|  | 0.5 pt | A large print application form is offered, but either is not directly available or requires more clicks to navigate to it. Partial credit is given if the state provides contact information for requesting a large-print form. | Arkansas has a large print form available, but applicant must contact the DHS office to obtain it. |
|  | 0 pt | The state does not mention a large print form on its website (e.g., no credit is given if this information is only found in the PDF application). | Kentucky does not receive credit for providing large print upon request, because information about this option is only found in its PDF application. |
